# Exploring the potential for collaborative use of an app-based platform for n-of-1 trials among healthcare professionals that treat patients with insomnia

**DOI:** 10.1101/2020.01.31.20019927

**Authors:** Jason R. Bobe, Jessica K. De Freitas, Benjamin S. Glicksberg

## Abstract

**Background:** N-of-1 trials are single patient, multiple crossover, comparative effectiveness experiments. Despite their rating as “level 1” evidence, they are not routinely used in clinical medicine to evaluate the effectiveness of treatments.

**Objective:** We explored the potential for implementing a mobile app-based n-of-1 trial platform for collaborative use by clinicians and patients to support data-driven decisions around the treatment of insomnia.

**Methods:** A survey assessing awareness and utilization of n-of-1 trials was administered to healthcare professionals that frequently treat patients with insomnia at the Icahn School of Medicine at Mount Sinai in New York City. 1M electronic health records were analyzed to evaluate evidence for a comorbid relationship between insomnia and dementia or Alzheimer’s disease among a patient population that may benefit from n-of-1 trials for the selection of optimal sleep treatments.

**Results:** A total of 45 healthcare professionals completed the survey and were included in the analysis. We found that 64% of healthcare professionals surveyed had not heard of n-of-1 trials. After a brief description of these methods, 75% of healthcare professionals reported that they are likely or highly likely to use an app-based n-of-1 trial at least once in the next year if the service were free and easy to offer to their patients.

**Conclusions:** An app-based n-of-1 trials platform might be a valuable tool for clinicians and patients to identify the best treatments for insomnia. Educational interventions that raise awareness and provide training are also likely necessary. The electronic health record (EHR) may help identify eligible patients.

## 1 Introduction

Healthcare professionals routinely practice individualized care. They design treatment plans based on unique patient characteristics and clinical presentation, consider various levels of evidence for treatment efficacy, and help patients weigh the risks of side effects and other potential treatment burdens and trade-offs. While there is widespread agreement that we should not expect most treatments to work uniformly across most populations, the systematic evaluation of treatment effect remains a challenge for healthcare professionals and patients [1]. Healthcare professionals who practice “evidence-based medicine” generally use comparisons of means and proportions between large groups of patients (e.g. from clinical trials), but are intuitively aware that there exists large heterogeneity of effects for many disease processes and interventions.(possible references). N-of-1 trials create an opportunity in some contexts for healthcare professionals and patients to individualized treatment selection in a more systematic way. They are designed to help both parties make objective, data-driven treatment choices.

### 1.1 What are n-of-1 trials?

In clinical medicine, N-of-1 trials are used as a decision support tool to inform individualized treatment selection [2]. The Oxford Centre for Evidence-Based Medicine ranks n-of-1 trials as “level 1” evidence for determining whether a treatment benefits a patient [3]. N-of-1 trials are also viewed as a tool to enhance patient-centered care, insofar as the patient may be involved in the selection of treatments to compare, the selection of outcomes to measure, and the selection of the treatment to continue at the end of the trial [4]. Typically in an n-of-1 trial, a single patient completes a baseline period without any treatments, then alternates between two treatments in a sequence (i.e. “multiple crossover”) [5,6]. Where feasible, treatments may be blinded or placebo-controlled. Outcomes are measured during baseline and each treatment period. At the end of the trial, outcome measurements for each treatment are compared and a treatment is selected. N-of-1 trials may also be deployed to answer other common treatment investigations, such as whether to begin a treatment, proper dosing [7], disease-related nutrition recommendations [8–10], assessing treatment response in people with characteristics (e.g. rare genetic variants) not studied in randomized controlled trials of approved medications [11–13], and deprescribing [14], among others.

To be sure, n-of-1 trials are not useful in every context. They work best in patients with chronic or stable conditions. Non-curative treatments with rapid onset and rapid washout are ideal candidates for n-of-1 trials, whereas treatments with cumulative effectiveness (e.g. some antidepressants) or treatments that disrupt the nature of the underlying condition (e.g. surgery) are not. N-of-1 trials are particularly relevant in contexts where evidence for treatment efficacy is weak or where treatment effects are known to be heterogeneous across populations and among individuals [5].

### 1.2 Chronic insomnia: a testbed for implementation of n-of-1 trials

There are many therapeutic contexts where n-of-1 trials are able to serve unmet patient needs. Chronic insomnia is a good target disorder because there is a high prevalence of affected individuals in the general population and also across distinct clinical populations, including Alzheimer’s disease (AD), Parkinson’s disease (PD), attention deficit hyperactivity disorder (ADHD), pregnancy, post-treatment cancer patients, and many others.

N-of-1 trials have been implemented or are currently underway in several populations with insomnia. Coxeter et al. reported the effectiveness of valerian versus placebo in 24 patients with chronic insomnia [15]. Punja et al. have described a protocol to assess the effectiveness of melatonin in children (ages 6-17) taking medications for ADHD [16]. Nikles et al. have described a protocol to assess the effectiveness of melatonin in patients with PD [17].

Many patient populations have insomnia. It is one of the most common mental disorders [18]. Patients with insomnia have significant interest in finding the most effective treatment due to the negative impact on quality-of-life for patients, family and caregivers [19,20]. Poor sleep may also interact with recovery or progression of some diseases, including cardiovascular disease, depression, diabetes, AD, among others [21–25]. Estimates of the prevalence of insomnia in the general population vary in part due to the definition of insomnia used. The American Academy of Sleep Medicine (AASM) estimates that 33-50% of adults experience symptoms of insomnia and 10-15% of adults experience insomnia disorders that disrupt daily functioning [26].

The International Classification of Sleep Disorders, 3rd Edition (ICSD-3) includes seven categories of sleep disorders, including insomnia. The category of insomnia has three sub-types: chronic insomnia disorder, short-term insomnia disorder, and other insomnia disorder. The criteria for diagnosing chronic insomnia specify that a person must report a problem with sleep initiation or maintenance while having an adequate opportunity and environment to sleep. Symptoms must also cause clinically significant functional distress during the daytime; be present for at least three nights per week for at least three months. Chronic insomnia is likely a heterogeneous disorder [27]. Brain imaging studies, for example, have not found consistent patterns across patients with chronic insomnia [28].

There is limited efficacy data available for insomnia treatments in many patient populations, so decisional conflict is common. The American Academy of Physicians recommends cognitive behavioral therapy for insomnia (CBT-I) as the first line therapy in adults with chronic insomnia [29]. In a systematic review of randomized controlled trials (RCTs) that compared CBT-I and medications for the treatment of insomnia, evidence supports the notion that CBT-I is better than medications in some contexts [30]. However, among patients that choose to pursue CBT-I, around 20-30% fail to respond [31]. Furthermore, many people do not choose CBT-I because of several treatment-related burdens, including limited access to trained healthcare professionals, weekly therapy appointments, and out-of-pocket costs [32,33]. There is widespread use of pharmacological interventions and over-the-counter sleep aids. Around 20% of U.S. adults use prescribed or over-the-counter (OTC) sleeping medications each month [34]. While commonly used, many OTC sleep treatments have limited efficacy and safety data [35]. Furthermore, some sleep medications that are commonly used in younger populations for sleep problems are, for example, potentially inappropriate for use in older populations (e.g. hypnotics or Z drugs) [36].

For n-of-1 trials to be an effective tool for patients and healthcare professionals, the design of these trials should incorporate the precise needs of the populations they intend to serve. Next, we describe one patient population where chronic sleep disturbance is a salient issue: individuals at risk for AD with chronic sleep problems.

### 1.3 Alzheimer’s and sleep disturbance

AD is a critical public health concern with an estimated 131 million people worldwide predicted to be affected by the year 2050 [37]. More than six million older Americans have AD [38,39] and the annual incidence of the disease is estimated to rise to one million cases by 2050, which is more than double the current incidence [40]. Research into possible preventive measures is an urgent need, especially for people at risk due to family history or genetic predisposition. Primary prevention of AD is currently seen as the most effective strategy at reducing the prevalence of AD over the next several decades [39]. In recent years, sleep has been identified as a potential modifiable risk factor for prevention or delay of AD [41].

The connection between poor sleep and negative outcomes among the elderly population has long been established. In the 1990s, a prospective study of elderly people living in New York City found that insomnia among males was the strongest predictor of mortality and placement in a nursing home, exceeding many other factors analyzed, including age, income, health, and activities of daily living (ADL) [42]. Another prospective study found that older women with sleep disturbance were more likely to get dementia or mild cognitive impairment (MCI) [43,44].

In 2017, a meta-analysis of 27 observational studies found that poor sleep might explain around ∼15% of cases of cognitive impairment or AD [45]. They observed that people with sleep problems had a 1.55, 1.65, and 3.78 times higher risk for AD, cognitive impairment, and preclinical AD than individuals without sleep problems, respectively [45]. They also examined the relative risks related to specific sleep problems, including short and long sleep duration (1.86), poor sleep quality (1.62), circadian rhythm abnormality (1.38), and insomnia (1.38) [45].

The conventional wisdom is that changes in the brain due to AD causes sleep disturbance. Indeed, the prevalence of sleep problems among patients with AD range from 25-50% [46]. Recent evidence suggests causality might also work in the other direction: sleep disturbance as a risk factor for developing AD [24,41,47,48]. Several recent studies provide evidence of a plausible mechanism: increased amyloid-β production during periods of sleep deprivation. A single night of sleep deprivation leads to elevated amyloid-β-42 (Aβ42) in the cerebrospinal fluid of healthy middle-aged men [49]. In a small, 3-arm, crossover RCT researchers found that sleep deprivation increased overnight amyloid-β (−38, −40, −42) levels by 25–30%, while regular sleep or sedated sleep showed no change in Aβ levels [50]. The connection between sleep disturbance and risk for AD remains unproven and other plausible mechanisms have been proposed [51].

With the emerging evidence around the importance of sleep, people with a family history of Alzheimer’s disease and those with genetic risk factors (e.g. APOE-e4) are likely to be motivated to find the most effective interventions to address chronic sleep problems. Indeed, the REVEAL Study observed healthy behavior change among patients that received genetic risk information for Alzheimer’s disease, even in the absence of any proven approaches to prevention [52]. This group of individuals are likely to meet many of the criteria identified by clinicians in qualitative interviews about ideal patients for n-of-1 trials: “proactive, cognitively intact, reliable, motivated, and engaged in a trusting physician–patient relationship” [53].

### 1.4 Key questions for the current study

The purpose of this study is to collect preliminary evidence to inform potential routes for the implementation of an app-based n-of-1 trial platform, called the N1 app. While this platform may be adapted to inform the optimal selection of treatments for a variety of disorders and wellness-related goals, patients with insomnia have been identified as a potential population to target due to the prevalence of the condition, limited evidence for efficacy of treatments across some patient populations, the possibility to incorporate wearable devices for the passive collection of outcome measures [54], among others.

In this study we aim to assess the familiarity and experience with n-of-1 trials in a sample of healthcare workers that frequently care for patients with insomnia. Further, we examine the characteristics of a hospital patient population that may stand to benefit from n-of-1 trials for the selection of optimal sleep treatments. One hypothesis is that healthcare professionals are currently familiar with, but do not regularly use n-of-1 trials. A second hypothesis is that data mining of Electronic Health Records (EHR) in a hospital, health system, or health plan could be a viable route for n-of-1 trial patient recruitment, which we explore through the assessment of the comorbidity relationship between insomnia and dementia.

## 2 METHODS

### 2.1 Survey

#### 2.1.1 Instrument

The Office for the Protection of Human Subjects at Mount Sinai approved a protocol to administer a voluntary, anonymous survey to healthcare professionals. The exploratory, cross-sectional survey administered has three sections (a) sociodemographics (b) experience and satisfaction with the treatment of patients with insomnia and (c) awareness and utilization of n-of-1 trials (see Datasheet S1).The survey was administered online through REDCap (Research Electronic Data Capture) hosted by the Icahn School of Medicine at Mount Sinai [55].

#### 2.1.2 Sample and recruitment

We chose to recruit a sample of clinicians and nurse practitioners through the Department of Psychiatry due to the regularity with which these healthcare professionals treat patients with insomnia. We sent messages through the department mailing list and advertised at bimonthly Grand Rounds events during October and November of 2019.

#### 2.1.3 Statistical analysis

We primarily use standard descriptive statistics to summarize the survey results. We further assessed differences of participants awareness of n-of-1 using two-sided Chi-squared test for categorical variables and a t-test for continuous variables. Using the same methods, we also assessed associations for willingness to use a n-of-1 digital service, which we binarized due to low sample size and imbalanced responses into “More likely” (consisting of “Strongly agree” and “Agree” responses) and “Less likely” (consisting of “Neutral”, “Disagree”, and Strongly disagree” responses).

### 2.2 Electronic Health Records Analysis

#### 2.2.1 Clinical Cohort

We utilize clinical data from the EHRs of the Mount Sinai Hospital (MSH). MSH is an urban, tertiary care hospital located on the Upper East Side of Manhattan in New York City. The clinical data contained within the EHRs consists of structured data of patient information such as encounters, disease diagnoses, lab test results, vital signs, medication prescriptions, and procedures among others.

#### 2.2.2 Phenotyping process and cohort breakdown

For the current study, we restrict our research cohort to individuals with at least one recorded clinical feature, who were at least 70 years old, with a minimum of five years of medical history in our system, and ICD codes for both insomnia and AD or dementia (See Datasheet S2), leaving 2632 unique patients for subsequent analyses (see Figure S1). This filtering procedure allowed for more robust phenotyping by prioritizing individuals 65 and older that are more likely to be regular patients in the MSH system (i.e., currently 70 years or older with at least five years of prior history and at least one diagnostic code). Due to HIPAA requirements, the ages of patients within the research cohort are right-censored at age 90. The mean age of the cohort is 84.3 ± 12.5 (std). The self-reported sex breakdown of the cohort is 71.1% female, 28.8% male, and 0.04% not available. The self-reported race breakdown of the cohort is: 50.8% Caucasian (White), 30.4% African American (Black), 12.8% Other, 6.6% Unknown, and 2.7% Asian. The self-reported ethnicity breakdown of the cohort is: 43.9% Hispanic/Latino, 25.5% non-Hispanic/Latino, and 31.9% Unknown.

#### 2.2.3 Enrichment analysis

We used a chi-squared test to evaluate the relationship between insomnia and AD or dementia. Our null hypothesis is that the diseases are independent and they have no significant comorbidity relationship. Specifically, we tabulated the number of individuals that pass the above filtering criteria that have both AD or dementia diagnosis and an insomnia diagnosis, compared to those with one or the other, and to those with neither.

### 2.3 Ethics approval

This study has been approved by the Institutional Review Board at the Icahn School of Medicine at Mount Sinai (IRB-18-00789 & IRB-19-02369)

## 3 RESULTS

### 3.1 Survey Results

#### 3.1.1 Sample characteristics

A total of 66 participants consented to the survey, 49 participants completed the survey and the responses from 45 participants were included in the analysis (see Figure S2). This sample of 45 healthcare professionals from a large, urban hospital are mostly practicing clinicians (88.9%), affiliated with the Department of Psychiatry (88.9%), in their first or second decade of clinical practice (64.4%), with a primary specialty of Psychiatry and Neurology (93.3%) (see Table 1).

**Table 1:** Sociodemographics of the sample of healthcare professionals surveyed

| Category | Variable | n | % |
| --- | --- | --- | --- |
| **Age (mean +/- SD)** | 48.4 +/- 16.5 | 45 | 100.0 |
| **Sex** | Female | 26 | 57.8 |
|  | Male | 18 | 40.0 |
|  | Not reported | 1 | 2.2 |
| **Race** | Asian | 3 | 6.7 |
|  | Black or African American | 2 | 4.4 |
|  | White | 34 | 75.6 |
|  | Multiple | 3 | 6.7 |
|  | Unknown / Not reported | 3 | 6.7 |
| **Ethnicity** | Hispanic/Latino | 3 | 6.7 |
|  | Not Hispanic/Latino | 41 | 91 |
|  | Unknown/Not reported | 1 | 2.2 |
| **Clinical degree(s)** | MD | 39 | 86.7 |
|  | NP | 5 | 11.1 |
|  | MD/NP | 1 | 2.2 |
| **Years of clinical practice** | 0-10 | 20 | 44.4 |
|  | 11-20 | 9 | 20.0 |
|  | 21-30 | 7 | 15.6 |
|  | 31-40 | 5 | 11.1 |
|  | >40 | 3 | 6.7 |
|  | Not reported | 1 | 2.2 |
| **Department(s)** | Psychiatry | 43 | 95.6 |
|  | Psychiatry and Anesthesiology | 1 | 2.2 |
|  | Not reported | 1 | 2.2 |
| **Primary Specialty** | Psychiatry and Neurology | 42 | 93.3 |
|  | Other | 1 | 2.2 |
|  | Not reported | 2 | 4.4 |
| **Total** | - | 45 | 100.0 |

#### 3.1.2 Experience and satisfaction of sample with current treatments for patients with insomnia

Most healthcare professionals surveyed frequently see patients with insomnia in their clinical practice with 88.9% reporting daily or weekly encounters. A majority of participants also expressed their dissatisfaction with available treatment options for their patients with insomnia, with 55.6% disagreeing or strongly disagreeing with the statement: “I am satisfied with the available treatment options for my patients with insomnia.” Participants also perceived their patients as being dissatisfied with available treatment options, with 62.2% disagreeing or strongly disagreeing with the statement: “My patients are satisfied with available treatment options for insomnia” (see Table 2).

**Table 2.**
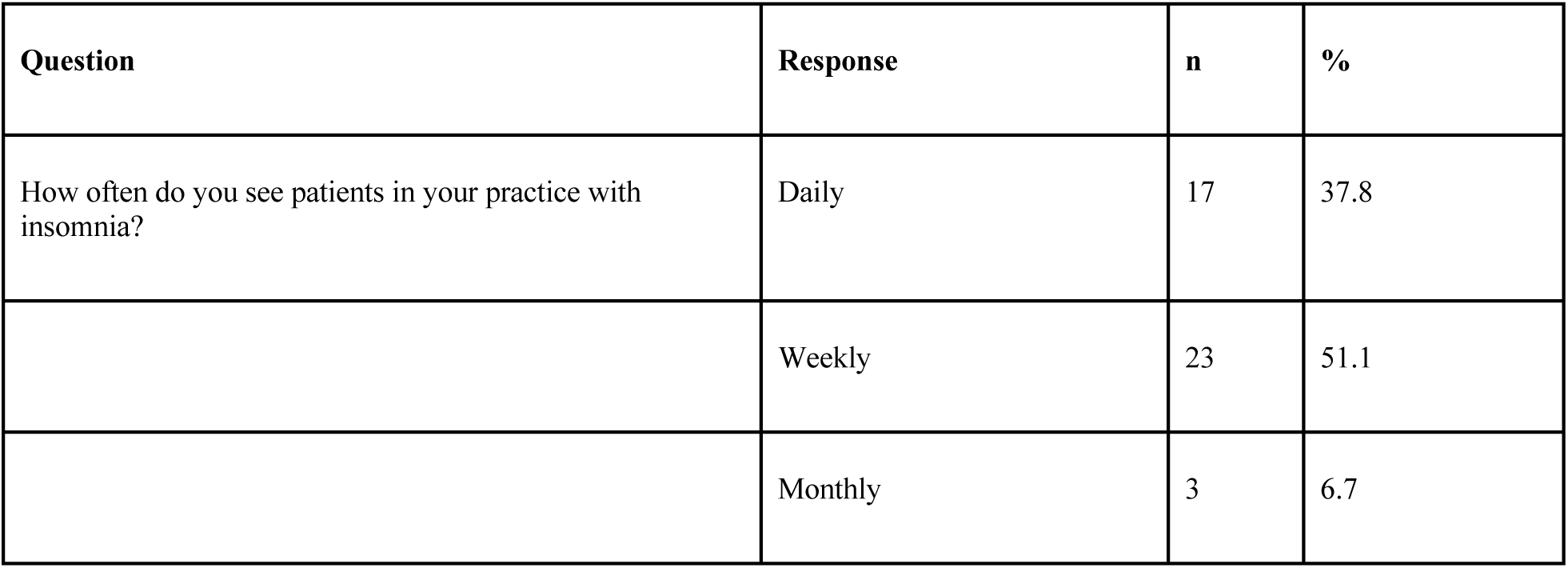
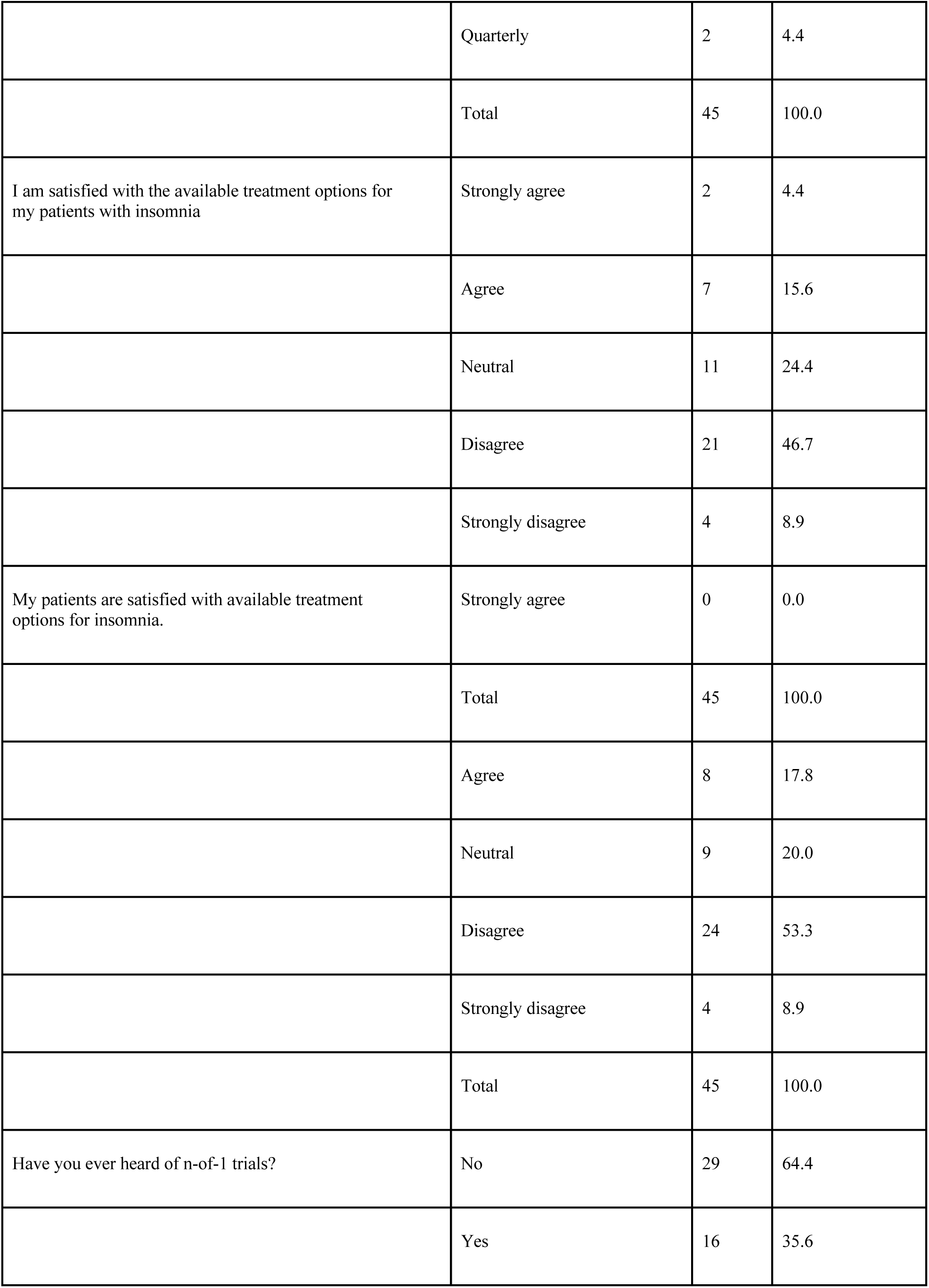
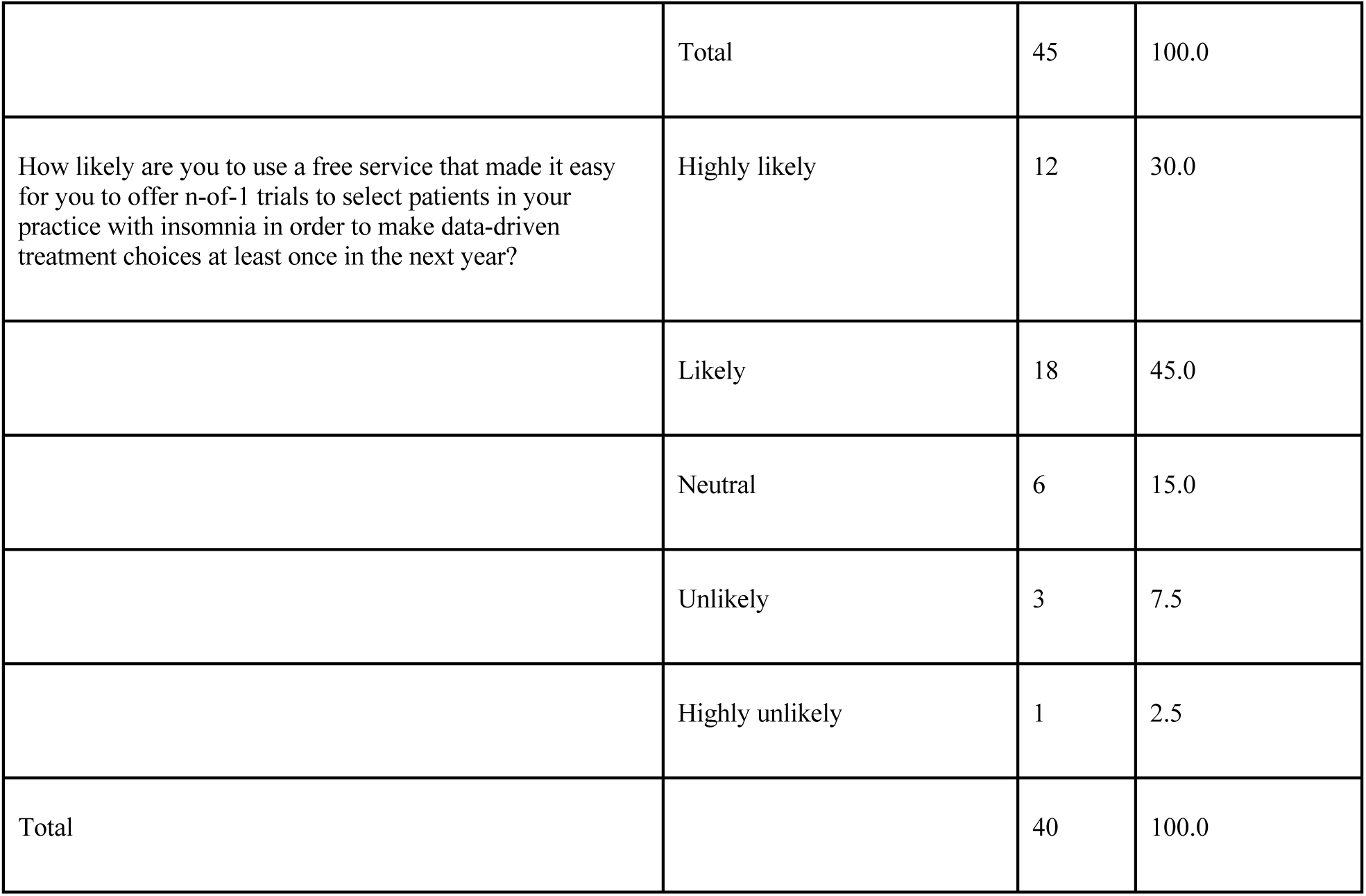
Summary of survey results from a sample of healthcare professionals on their experience and satisfaction with current treatments for insomnia

| Question | Response | n | % |
| --- | --- | --- | --- |
| How often do you see patients in your practice with insomnia? | Daily | 17 | 37.8 |
|  | Weekly | 23 | 51.1 |
|  | Monthly | 3 | 6.7 |
|  | Quarterly | 2 | 4.4 |
|  | Total | 45 | 100.0 |
| I am satisfied with the available treatment options for my patients with insomnia | Strongly agree | 2 | 4.4 |
|  | Agree | 7 | 15.6 |
|  | Neutral | 11 | 24.4 |
|  | Disagree | 21 | 46.7 |
|  | Strongly disagree | 4 | 8.9 |
| My patients are satisfied with available treatment options for insomnia. | Strongly agree | 0 | 0.0 |
|  | Total | 45 | 100.0 |
|  | Agree | 8 | 17.8 |
|  | Neutral | 9 | 20.0 |
|  | Disagree | 24 | 53.3 |
|  | Strongly disagree | 4 | 8.9 |
|  | Total | 45 | 100.0 |
| Have you ever heard of n-of-1 trials? | No | 29 | 64.4 |
|  | Yes | 16 | 35.6 |
|  | Total | 45 | 100.0 |
| How likely are you to use a free service that made it easy for you to offer n-of-1 trials to select patients in your practice with insomnia in order to make data-driven treatment choices at least once in the next year? | Highly likely | 12 | 30.0 |
|  | Likely | 18 | 45.0 |
|  | Neutral | 6 | 15.0 |
|  | Unlikely | 3 | 7.5 |
|  | Highly unlikely | 1 | 2.5 |
| Total |  | 40 | 100.0 |

#### 3.1.3 Awareness and use of n-of-1 trials

Most participants surveyed were unfamiliar with the concept of n-of-1 trials, with 64.4% reporting that they had never heard of them before. Following this survey question, participants were presented with a short description of n-of-1 trials that also included a screenshot of the N1 app, a smartphone based n-of-1 trial platform developed at the Icahn School of Medicine (see Data sheet S1) [56,57]. They were asked to consider the scenario that “a free service [existed] that made it easy for you to offer n-of-1 trials to select patients in your practice with insomnia. The patient would conduct the mobile app-based trial at home. At the conclusion of the trial, the analyzed results would be available to you and the patient to review together.” Excluding 5 participants that did not respond to this question, 75% of participants (i.e. 30 of 40) reported that they were either “likely” or “highly likely” to “use a service like this to make data-driven treatment choices at least once in the next year.”

Of the 16 healthcare professionals that reported that they were aware of n-of-1 trials, 3 reported that they had previously used an n-of-1 trial in the treatment of their patients. For the remaining 11 participants that were aware of n-of-1 trials, had never used them in their clinical practice, and reported a primary reason for the lack of adoption, 45.4% cited inadequate training in n-of-1 trial design.

We further assessed the relationship between various questionnaire features and having heard of n-of-1 trials (n=45). We found no significant relationship between having heard of n-of-1 trials and age (p = 0.54, t = −0.61), sex (p = 0.09, X-squared = 2.91), race (p = 0.10, X-squared = 7.92), ethnicity (p = 0.75, X-squared = 0.58), clinical degree (p = 0.84, X-squared = 0.04; one individual with both degrees not included), number of years practiced (p = 0.29, X-squared = 5.02), frequency of seeing patients with insomnia (p = 0.50, X-squared = 2.36), satisfaction of current treatments for insomnia (p = 0.23, X-squared = 5.65), or perceived patient satisfaction of current treatments for insomnia (p = 0.80, X-squared = 0.99).

We also assessed the relationship between various questionnaire features and willingness to use a n-of-1 digital service app (n=40; binarized response). We found no significant relationship between willingness to use a digital app service and age (p = 0.22, t = −1.27), sex (p =0.23, X-squared =1.45), race (p =0.54, X-squared = 3.13), ethnicity (p = 0.20, X-squared = 3.26), clinical degree (p = 0.81, X-squared = 0.06; one individual with both degrees not included), number of years practiced (p = 0.67, X-squared = 2.36), frequency of seeing patients with insomnia (p = 0.64, X-squared = 1.68), satisfaction of current treatments for insomnia (p = 0.21, X-squared = 5.87), or perceived patient satisfaction of current treatments for insomnia (p = 0.82, X-squared = 0.93).

### 3.2 Electronic Health Records clinical comorbidity analysis

In the EHR analysis, our goal was to discern whether an enriched comorbidity relationship between AD or dementia and insomnia was present in the MSH patient population. We present the breakdown of patients by ICD code for each disease category in Figure 1 (A and B). We further visualize the distribution and frequency of ICD codes between both disease categories in Figure 1C.

**Figure 1:**
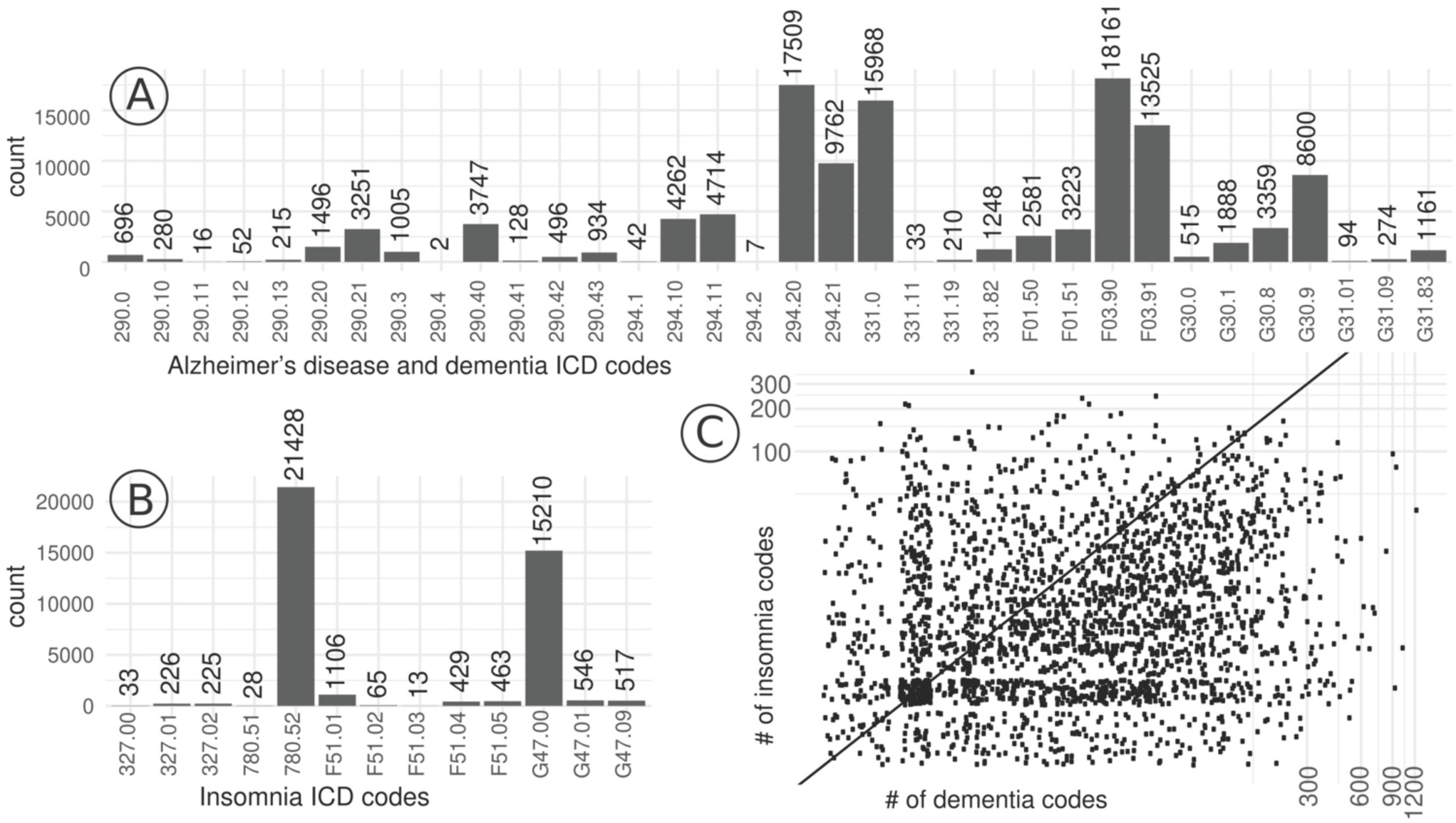
The breakdown of patients by each ICD-9-CM and ICD-10-CM code of each category, specifically for A) Alzheimer’s disease or dementia and B) insomnia. Note that there might be patients that have more than one associated code. C) a scatter plot visualizing the distribution and frequency of patients by number of ICD codes of each disease category. Here, we display the number of ICD codes associated for each patient for insomnia codes on the y-axis and AD or dementia codes on the x-axis.

The null hypothesis was that the two diseases were independent from one another. We performed a chi-square assessment for the rates of individuals that passed our eligibility criteria (see Figure 1) had instances of both diseases compared to one or the other against a background of those with neither (see Table 3). From this analysis, we obtained a chi-squared test statistic of 2978 which corresponds to a statistically significant p-value of < 2.2 e-16. Odds ratio was used to quantify the strength of the relationship between insomnia and Alzheimer’s/dementia. We obtained a relative risk of 3.01 and an odds ratio of 3.57, with a 95% confidence interval of 3.40-3.75.

**Table 3:** A contingency table tabulating the number of patients in the Mount Sinai Hospital Electronic Health Records that pass our filtering criteria with Alzheimer’s disease (AD) or dementia and insomnia.

|  | AD/Dementia | No AD/dementia |
| --- | --- | --- |
| Insomnia | 2514 | 8997 |
| No insomnia | 10724 | 137030 |

## 4 DISCUSSION

The N1 app aims to facilitate the design, administration, and analysis of n-of-1 trials [56–58]. Individuals with insomnia or other chronic sleep disturbance issues are a population that may benefit from access to n-of-1 trials for data-driven treatment selection. While these multi-crossover, comparative effectiveness trials have been in use for decades, awareness and adoption by healthcare professionals continues to face challenges, as our survey further indicates. Yet, we are encouraged that the healthcare professionals sampled did also report substantial receptivity to future use of app-based n-of-1 trials to inform the treatment of patients with insomnia provided there were a service that made such trials free and easy to implement. The lack of awareness of n-of-1 trials coupled with receptivity to their use suggests that educational interventions may address at a current barrier to wider utilization of n-of-1 trials.

Historically, several centers have been established to support the implementation of these trials for local clinicians as a service [7,53,59]. One center currently operates at the University of Queensland in Australia with a focus on sleep [60]. While these centers have documented many cases where patients and clinicians were aided in the selection of treatments, the centers are often experiments themselves that last as long as funding permits their operation, for example. For a time in the United States, there was a Current Procedural Terminology (CPT) code for “personalized medicine tests” that suggested a route to reimbursement for the effort required to design, administer and analyze an n-of-1 trial [4]. Writing in 2008, Kravitz et al. suggested that one alternative to the model where n-of-1 trials take root and gain traction primarily through academic clinical centers is the possibility that they “cast off some of their academic trappings and focus on appealing to what patients want and need” [4].

One of the major challenges with prospective studies, including n-of-1 trials, is cohort identification and recruitment. A benefit of working within a health system is the possibility of accessing retrospective patient data within the EHR. Of course, there are plenty of challenges of working with such data (e.g., bias and missingness) but the EHR also offers countless opportunities for research and a promising resource for patient engagement [61]. For instance, study recruitment via EHRs and patient portals are an emerging trend for pragmatic trials [62]. In our EHR analysis, we found a significant comorbidity enrichment between sleep disturbances, specifically insomnia, and AD or dementia. This finding is encouraging as it replicates a previously established association and reinforces the notion of using the EHR to aid in the construction of cohorts with specific features for research and recruitment.

While healthcare practitioners take into consideration each patient’s unique characteristics and strive to use the most up-to-date information to make informed therapeutic recommendations, there will always be some variability and uncertainty in outcomes. Leveraging n-of-1 trials as self-contained experiments can quantify individual outcomes and can better optimize treatment selection in some contexts. We believe growth in the adoption of n-of-1 trials will enhance the precision of treatment selection for many individuals. App-based platforms offer the potential to reduce some barriers, but other challenges still remain.

## Data Availability

The survey and electronic health record datasets for this manuscript are not publicly available because of privacy considerations of patients and clinical practitioners.

## 5 Acknowledgments

This study is funded by the Icahn School of Medicine at Mount Sinai and a generous gift from Julian Salisbury. Thanks to Drs. Richard Kravitz MD (UC Davis) and Andrew Avins MD (Kaiser Permanente) for providing feedback on the survey instrument and the manuscript, and also thanks to Alex Charney MD PhD (Mount Sinai) for providing feedback on the survey instrument. The authors declare that the research was conducted in the absence of any commercial or financial relationships that could be construed as a potential conflict of interest.

## 6 Author contributions

All authors listed have made a substantial, direct and intellectual contribution to the work, and approved it for publication.

## References

1. Lillie EO, Patay B, Diamant J, Issell B, Topol EJ, Schork NJ. The n-of-1 clinical trial: the ultimate strategy for individualizing medicine? Per Med. 2011;8: 161–173. doi:10.2217/pme.11.7

2. Samuel J, Holder T, Molony D. N-of-1 Trials as a Decision Support Tool in Clinical Practice: A Protocol for a Systematic Literature Review and Narrative Synthesis. Healthcare (Basel). 2019;7. doi:10.3390/healthcare7040136

3. OCEBM Levels of Evidence - CEBM. In: CEBM [Internet]. 1 May 2016 [cited 8 Oct 2019]. Available: https://www.cebm.net/2016/05/ocebm-levels-of-evidence/

4. Kravitz RL, Duan N, Niedzinski EJ, Hay MC, Subramanian SK, Weisner TS. What ever happened to N-of-1 trials? Insiders’ perspectives and a look to the future. Milbank Q. 2008;86: 533–555. doi:10.1111/j.1468-0009.2008.00533.x

5. Mitchell G, Nikles J, editors. The Essential Guide to N-of-1 Trials in Health. Springer; 2015. Available: https://www.springer.com/gp/book/9789401771993

6. Kravitz RL, Duan N, eds, and the DEcIDE Methods Center N-of-1 Guidance Panel (Duan N, Eslick I, Gabler NB, Kaplan HC, Kravitz RL, Larson EB, Pace WD, Schmid CH, Sim I, Vohra S). Design and Implementation of N-of-1 Trials: A User’s Guide. Rockville, MD: AHRQ; 2014 Jan. Report No.: Publication No. 13(14)-EHC122-EF. Available: https://effectivehealthcare.ahrq.gov/products/n-1-trials/research-2014-5

7. Guyatt GH, Keller JL, Jaeschke R, Rosenbloom D, Adachi JD, Newhouse MT. The n-of-1 randomized controlled trial: clinical usefulness. Our three-year experience. Ann Intern Med. 1990;112: 293–299. doi:10.7326/0003-4819-112-4-293

8. Kaplan HC, Opipari-Arrigan L, Schmid CH, Schuler CL, Saeed S, Braly KL, et al. Evaluating the Comparative Effectiveness of Two Diets in Pediatric Inflammatory Bowel Disease: A Study Protocol for a Series of N-of-1 Trials. Healthcare (Basel). 2019;7. doi:10.3390/healthcare7040129

9. Liu L, Zhang Y, Wei J, Chen Z, Yu J. A Pilot Study of Amino Acids in Unresectable Non-Small-Cell Lung Cancer Patients During Chemotherapy: A Randomized Serial N-of-1 Trials Design. Nutr Cancer. 2019;71: 399–408. doi:10.1080/01635581.2018.1515962

10. Soldevila-Domenech N, Boronat A, Langohr K, de la Torre R. N-of-1 Clinical Trials in Nutritional Interventions Directed at Improving Cognitive Function. Front Nutr. 2019;6: 110. doi:10.3389/fnut.2019.00110

11. Magaret AS, Mayer-Hamblett N, VanDevanter D. Expanding access to CFTR modulators for rare mutations: The utility of n-of-1 trials. J Cyst Fibros. 2019. doi:10.1016/j.jcf.2019.11.011

12. Nick JA, St Clair C, Jones MC, Lan L, Higgins M, VX12-770-113 Study Team. Ivacaftor in cystic fibrosis with residual function: Lung function results from an N-of-1 study. J Cyst Fibros. 2019. doi:10.1016/j.jcf.2019.09.013

13. Griggs RC, Batshaw M, Dunkle M, Gopal-Srivastava R, Kaye E, Krischer J, et al. Clinical research for rare disease: opportunities, challenges, and solutions. Mol Genet Metab. 2009;96: 20–26. doi:10.1016/j.ymgme.2008.10.003

14. Clough AJ, Hilmer SN, Naismith SL, Gnjidic D. The Feasibility of Using N-Of-1 Trials to Investigate Deprescribing in Older Adults with Dementia: A Pilot Study. Healthcare (Basel). 2019;7. doi:10.3390/healthcare7040161

15. Coxeter PD, Schluter PJ, Eastwood HL, Nikles CJ, Glasziou PP. Valerian does not appear to reduce symptoms for patients with chronic insomnia in general practice using a series of randomised n-of-1 trials. Complement Ther Med. 2003;11: 215–222. doi:10.1016/s0965-299(03)00122-5

16. Punja S, Nikles CJ, Senior H, Mitchell G, Schmid CH, Heussler H, et al. Melatonin in Youth: N-of-1 trials in a stimulant-treated ADHD Population (MYNAP): study protocol for a randomized controlled trial. Trials. 2016;17: 375. doi:10.1186/s13063-016-1499-6

17. Nikles J, O’Sullivan JD, Mitchell GK, Smith SS, McGree JM, Senior H, et al. Protocol: Using N-of-1 tests to identify responders to melatonin for sleep disturbance in Parkinson’s disease. Contemp Clin Trials Commun. 2019;15: 100397. doi:10.1016/j.conctc.2019.100397

18. Wittchen HU, Jacobi F, Rehm J, Gustavsson A, Svensson M, Jönsson B, et al. The size and burden of mental disorders and other disorders of the brain in Europe 2010. Eur Neuropsychopharmacol. 2011;21: 655–679. doi:10.1016/j.euroneuro.2011.07.018

19. Kyle SD, Morgan K, Espie CA. Insomnia and health-related quality of life. Sleep Med Rev. 2010;14: 69–82. doi:10.1016/j.smrv.2009.07.004

20. Edelson J, Byrnes J, Mitchell G, Heussler H, Melaku M, Nikles J. Protocol for a Longitudinal Study of Melatonin Therapy and Cost Effectiveness Analysis in Stimulant-Treated Children with ADHD and Insomnia: An N-of-1 Trial. Contemporary Clinical Trials Communications. 2020; 100530. doi:10.1016/j.conctc.2020.100530

21. Jansen PR, Watanabe K, Stringer S, Skene N, Bryois J, Hammerschlag AR, et al. Genome-wide analysis of insomnia in 1,331,010 individuals identifies new risk loci and functional pathways. Nat Genet. 2019;51: 394–403. doi:10.1038/s41588-018-0333-3

22. Baglioni C, Battagliese G, Feige B, Spiegelhalder K, Nissen C, Voderholzer U, et al. Insomnia as a predictor of depression: a meta-analytic evaluation of longitudinal epidemiological studies. J Affect Disord. 2011;135: 10–19. doi:10.1016/j.jad.2011.01.011

23. Lane JM, Jones SE, Dashti HS, Wood AR, Aragam KG, van Hees VT, et al. Biological and clinical insights from genetics of insomnia symptoms. Nat Genet. 2019;51: 387–393. doi:10.1038/s41588-019-0361-7

24. Ju Y-ES, Lucey BP, Holtzman DM. Sleep and Alzheimer disease pathology--a bidirectional relationship. Nat Rev Neurol. 2014;10: 115–119. doi:10.1038/nrneurol.2013.269

25. Fan M, Sun D, Zhou T, Heianza Y, Lv J, Li L, et al. Sleep patterns, genetic susceptibility, and incident cardiovascular disease: a prospective study of 385 292 UK biobank participants. Eur Heart J. 2019. doi:10.1093/eurheartj/ehz849

26. Schutte-Rodin S, Broch L, Buysse D, Dorsey C, Sateia M. Clinical guideline for the evaluation and management of chronic insomnia in adults. J Clin Sleep Med. 2008;4: 487–504. Available: https://aasm.org/resources/clinicalguidelines/040515.pdf

27. Benjamins JS, Migliorati F, Dekker K, Wassing R, Moens S, Blanken TF, et al. Insomnia heterogeneity: Characteristics to consider for data-driven multivariate subtyping. Sleep Med Rev. 2017;36: 71–81. doi:10.1016/j.smrv.2016.10.005

28. Tahmasian M, Noori K, Samea F, Zarei M, Spiegelhalder K, Eickhoff SB, et al. A lack of consistent brain alterations in insomnia disorder: An activation likelihood estimation meta-analysis. Sleep Med Rev. 2018;42: 111–118. doi:10.1016/j.smrv.2018.07.004

29. Qaseem A, Kansagara D, Forciea MA, Cooke M, Denberg TD, Clinical Guidelines Committee of the American College of Physicians. Management of Chronic Insomnia Disorder in Adults: A Clinical Practice Guideline From the American College of Physicians. Ann Intern Med. 2016;165: 125–133. doi:10.7326/M15-2175

30. Mitchell MD, Gehrman P, Perlis M, Umscheid CA. Comparative effectiveness of cognitive behavioral therapy for insomnia: a systematic review. BMC Fam Pract. 2012;13: 40. doi:10.1186/1471-2296-13-40

31. Morin CM. Cognitive-behavioral approaches to the treatment of insomnia. J Clin Psychiatry. 2004;65 Suppl 16: 033–40. Available: https://www.ncbi.nlm.nih.gov/pubmed/15575803

32. Koffel E, Bramoweth AD, Ulmer CS. Increasing access to and utilization of cognitive behavioral therapy for insomnia (CBT-I): a narrative review. J Gen Intern Med. 2018;33: 955–962. doi:10.1007/s11606-018-4390-1

33. Morin CM. Issues and challenges in implementing clinical practice guideline for the management of chronic insomnia. J Sleep Res. 2017;26: 673–674. doi:10.1111/jsr.12639

34. Bertisch SM, Herzig SJ, Winkelman JW, Buettner C. National use of prescription medications for insomnia: NHANES 1999-2010. Sleep. 2014;37: 343–349. doi:10.5665/sleep.3410

35. Meolie AL, Rosen C, Kristo D, Kohrman M, Gooneratne N, Aguillard RN, et al. Oral nonprescription treatment for insomnia: an evaluation of products with limited evidence. J Clin Sleep Med. 2005;1: 173–187. Available: https://www.ncbi.nlm.nih.gov/pubmed/17561634

36. By the 2019 American Geriatrics Society Beers Criteria® Update Expert Panel. American Geriatrics Society 2019 Updated AGS Beers Criteria® for Potentially Inappropriate Medication Use in Older Adults. J Am Geriatr Soc. 2019;67: 674–694. doi:10.1111/jgs.15767

37. Alzheimer’s Disease International. World Alzheimer Report 2016. 2016. Available: https://www.alz.co.uk/research/WorldAlzheimerReport2016.pdf

38. Alzheimer’s Association. 2013 Alzheimer’s disease facts and figures. Alzheimers Dement. 2013;9: 208–245. doi:10.1016/j.jalz.2013.02.003

39. Brookmeyer R, Abdalla N, Kawas CH, Corrada MM. Forecasting the prevalence of preclinical and clinical Alzheimer’s disease in the United States. Alzheimers Dement. 2018;14: 121–129. doi:10.1016/j.jalz.2017.10.009

40. Hebert LE, Beckett LA, Scherr PA, Evans DA. Annual incidence of Alzheimer disease in the United States projected to the years 2000 through 2050. Alzheimer Dis Assoc Disord. 2001;15: 169–173. Available: https://www.ncbi.nlm.nih.gov/pubmed/11723367

41. Cordone S, Annarumma L, Rossini PM, De Gennaro L. Sleep and β-Amyloid Deposition in Alzheimer Disease: Insights on Mechanisms and Possible Innovative Treatments. Front Pharmacol. 2019;10: 695. doi:10.3389/fphar.2019.00695

42. Pollak CP, Perlick D, Linsner JP, Wenston J, Hsieh F. Sleep problems in the community elderly as predictors of death and nursing home placement. J Community Health. 1990;15: 123–135. doi:10.1007/BF01321316

43. Tranah GJ, Blackwell T, Stone KL, Ancoli-Israel S, Paudel ML, Ensrud KE, et al. Circadian activity rhythms and risk of incident dementia and mild cognitive impairment in older women. Ann Neurol. 2011;70: 722–732. doi:10.1002/ana.22468

44. Yaffe K, Laffan AM, Harrison SL, Redline S, Spira AP, Ensrud KE, et al. Sleep-disordered breathing, hypoxia, and risk of mild cognitive impairment and dementia in older women. JAMA. 2011;306: 613–619. doi:10.1001/jama.2011.1115

45. Bubu OM, Brannick M, Mortimer J, Umasabor-Bubu O, Sebastião YV, Wen Y, et al. Sleep, Cognitive impairment, and Alzheimer’s disease: A Systematic Review and Meta-Analysis. Sleep. 2017;40. doi:10.1093/sleep/zsw032

46. Moran M, Lynch CA, Walsh C, Coen R, Coakley D, Lawlor BA. Sleep disturbance in mild to moderate Alzheimer’s disease. Sleep Med. 2005;6: 347–352. doi:10.1016/j.sleep.2004.12.005

47. Roh JH, Huang Y, Bero AW, Kasten T, Stewart FR, Bateman RJ, et al. Disruption of the sleep-wake cycle and diurnal fluctuation of β-amyloid in mice with Alzheimer’s disease pathology. Sci Transl Med. 2012;4: 150ra122. doi:10.1126/scitranslmed.3004291

48. Minakawa EN, Wada K, Nagai Y. Sleep Disturbance as a Potential Modifiable Risk Factor for Alzheimer’s Disease. Int J Mol Sci. 2019;20. doi:10.3390/ijms20040803

49. Ooms S, Overeem S, Besse K, Rikkert MO, Verbeek M, Claassen JAHR. Effect of 1 night of total sleep deprivation on cerebrospinal fluid β-amyloid 42 in healthy middle-aged men: a randomized clinical trial. JAMA Neurol. 2014;71: 971–977. doi:10.1001/jamaneurol.2014.1173

50. Lucey BP, Hicks TJ, McLeland JS, Toedebusch CD, Boyd J, Elbert DL, et al. Effect of sleep on overnight cerebrospinal fluid amyloid β kinetics. Ann Neurol. 2018;83: 197–204. doi:10.1002/ana.25117

51. Lyon L. Is an epidemic of sleeplessness increasing the incidence of Alzheimer’s disease? rain. 2019;142: e30. doi:10.1093/brain/awz087

52. Chao S, Roberts JS, Marteau TM, Silliman R, Cupples LA, Green RC. Health behavior changes after genetic risk assessment for Alzheimer disease: The REVEAL Study. Alzheimer Dis Assoc Disord. 2008;22: 94–97. doi:10.1097/WAD.0b013e31815a9dcc

53. Kravitz RL, Paterniti DA, Hay MC, Subramanian S, Dean DE, Weisner T, et al. Marketing therapeutic precision: Potential facilitators and barriers to adoption of n-of-1 trials. Contemp Clin Trials. 2009;30: 436–445. doi:10.1016/j.cct.2009.04.001

54. Walch O, Huang Y, Forger D, Goldstein C. Sleep stage prediction with raw acceleration and photoplethysmography heart rate data derived from a consumer wearable device. Sleep. 2019. doi:10.1093/sleep/zsz180

55. Harris PA, Taylor R, Thielke R, Payne J, Gonzalez N, Conde JG. Research electronic data capture (REDCap)--a metadata-driven methodology and workflow process for providing translational research informatics support. J Biomed Inform. 2009;42: 377–381. doi:10.1016/j.jbi.2008.08.010

56. Bobe JR, Buros J, Golden E, Johnson M, Jones M, Percha B, et al. Factors Associated With Trial Completion and Adherence in App-Based N-of-1 Trials: Protocol for a Randomized Trial Evaluating Study Duration, Notification Level, and Meaningful Engagement in the Brain Boost Study. JMIR Res Protoc. 2020;9: e16362. doi:10.2196/16362

57. N1app.org. n: N1 app [Internet]. [cited 19 Jan 2020]. Available: http://n1app.org

58. Golden E, Johnson M, Jones M, Viglizzo R, Bobe J, Zimmerman N. Measuring the effects of caffeine and L-theanine on cognitive performance: a protocol for self-directed, mobile n-of-1 studies. 2019. doi:10.31219/osf.io/scnx6

59. Larson EB, Ellsworth AJ, Oas J. Randomized clinical trials in single patients during a 2-year period. JAMA. 1993;270: 2708–2712. doi:10.1001/jama.1993.03510220064035

60. N-of-1 trials and sleep research - Centre for Clinical Research - University of Queensland. [cited 20 Jan 2020]. Available: https://clinical-research.centre.uq.edu.au/nikles-group

61. Glicksberg BS, Johnson KW, Dudley JT. The next generation of precision medicine: observational studies, electronic health records, biobanks and continuous monitoring. Hum Mol Genet. 2018;27: R56–R62. doi:10.1093/hmg/ddy114

62. Pfaff E, Lee A, Bradford R, Pae J, Potter C, Blue P, et al. Recruiting for a pragmatic trial using the electronic health record and patient portal: successes and lessons learned. J Am Med Inform Assoc. 2019;26: 44–49. doi:10.1093/jamia/ocy138

